# Kilohertz Transcranial Magnetic Perturbation (kTMP) for Upper Extremity Motor Recovery After Stroke: A Nonrandomized Pilot Trial

**DOI:** 10.64898/2026.10.05.26364594

**Authors:** Christina M Merrick, Feiyang Dai, Saumya Singh, Ian O’Kidhain, Philipp Reber, Cidnee Luu, Angel V Peterchev, Daniel Sheltraw, Richard B Ivry, Ludovica Labruna, Karunesh Ganguly

## Abstract

**Importance:** Upper extremity impairment is a leading cause of post-stroke disability, yet clinically approved neuromodulation therapies are currently limited by the risks and costs of surgical implantation. There is an unmet need for non-invasive brain stimulation.

**Objective:** To evaluate the feasibility, safety, and preliminary clinical efficacy of a novel form of non-invasive brain stimulation, kilohertz transcranial magnetic perturbation (kTMP), for upper limb motor recovery after stroke.

**Design:** A single-center, open-label trial was conducted between September 2025 and April 2026 and comprised a 4-week standard-of-care baseline period, 4 weeks of active treatment with kTMP combined with motor training, and a 4-week follow-up.

**Setting:** Academic research center.

**Participants:** Seven individuals with chronic stroke and upper extremity motor deficits. **Intervention:** Four weeks of kTMP (3 sessions/week), a brain stimulation approach using magnetic induction to generate subthreshold electric fields, paired with motor tasks.

**Main Outcomes:** Change in Fugl-Meyer Assessment for Upper Extremity (FMA-UE) scores across study periods (standard-of-care baseline, active treatment, follow up). Secondary outcomes included measures of movement smoothness (Spectral Arc Length, SPARC) and fragmentation (mean velocity peak count), and intervention tolerability ratings.

**Results:** Across 7 participants (mean [SD] age, 57.4 [8.7] years; 4 males; mean [SD] time poststroke, 3.5 [2.1] years). No device-related adverse events occurred, and the intervention was well-tolerated (participants rated sensation intensity at an average of less than 0.5 on a 0-10 scale). FMA-UE scores remained stable during the baseline period (mean change, +0.14 points; 95% CI, –1.21 to 1.50; adjusted P =.98), increased significantly following the kTMP-intervention (mean change, +7.00 points; 95% CI, 5.31 to 8.69; adjusted P =.047), and were sustained at the 4-week follow-up (mean change, –1.33 points; 95% CI, –2.42 to –0.25; adjusted P =.13). Movement smoothness improved significantly within 1 week of kTMP onset (mean change, 1.48 points; 95% CI, 0.49–2.47; adjusted P =.03); movement fragmentation showed a similar but nonsignificant trend.

**Conclusions and Relevance:** This pilot trial provides preliminary evidence that kTMP combined with motor tasks is safe, feasible, and may support upper extremity recovery. These pilot data warrant large, randomized controlled trials to evaluate efficacy in support of poststroke motor recovery.

**Trial Registration: ClinicalTrials.gov Identifier:** NCT06317194

**Key Points:** *Question:* Is kilohertz transcranial magnetic perturbation (kTMP) a safe, feasible, and effective non-invasive intervention for improving upper extremity motor recovery in chronic stroke?

*Findings:* In this single-center, open-label pilot trial of 7 participants with chronic stroke, upper extremity motor impairment (FMA-UE) remained stable during a 4-week baseline period and significantly improved (mean change, +7.0 points) following 4 weeks of active kTMP treatment combined with motor tasks, with gains sustained at 4-week follow-up. No device-related adverse events occurred.

*Meaning:* These preliminary findings suggest that kTMP is safe and feasible and may support upper extremity motor recovery after stroke, warranting further evaluation.

## Introduction

Poststroke upper extremity impairment remains a major cause of disability^1^. The only FDA-approved neuromodulation method for stroke recovery is vagus nerve stimulation. However, its efficacy is variable^2^ and requires surgical implantation^3^. While non-invasive brain stimulation (NIBS) methods avoid surgical risks, none are currently FDA approved for stroke recovery. Moreover, existing NIBS methods have important limitations. Transcranial electrical stimulation (tES) produces relatively weak intracranial electric fields because of current shunting and is further constrained by scalp discomfort at higher intensities^4^; Transcranial magnetic stimulation (TMS) utilizes pulsed stimulation^5^ and can result in scalp discomfort as well^6^.

To address these limitations, we developed kilohertz transcranial magnetic perturbation (kTMP)^7^, a new form of NIBS that leverages magnetic induction to safely deliver robust subthreshold cortical electric fields (up to 8 V/m) while enabling amplitude-modulated (AM) waveforms designed to engage endogenous low-frequency motor rhythms implicated in poststroke motor recovery^8^. Building on prior demonstrations of safety and tolerability across 433 sessions in 143 participants^9^, we evaluated a 3-Hz AM-kTMP protocol combined with motor training in individuals with chronic upper-limb impairments after stroke. Upper limb motor recovery was assessed using the Fugl-Meyer Assessment for Upper Extremity (FMA-UE), comparing a 4-week standard-of-care baseline period with a 4-week intervention period. We hypothesized that active kTMP would produce greater improvements in FMA-UE scores than the baseline period.

## Methods

This prospective pilot study was conducted between September 2025 and April 2026 at the University of California, Berkeley (UCB). The trial protocol is provided in Supplement 1. The study was approved by the UCB Institutional Review Board, and all participants provided written informed consent. A total of 41 individuals were screened for eligibility. Thirty-three did not meet eligibility criteria and 1 declined participation, yielding a final cohort of 7 participants (Supplement Figure 1).

The 7 participants had a mean (SD) age of 57.4 (8.7) years. Four were men; five had ischemic stroke and two had hemorrhagic stroke. Mean (SD) time since stroke was 3.5 (2.1) years and mean (SD) baseline Fugl-Meyer Assessment–Upper Extremity (FMA-UE) score was 34.1 (11.0) (Table 1). Key eligibility criteria included age 18 to 80 years, upper limb motor dysfunction resulting from a single ischemic or hemorrhagic stroke at least 6 months before enrollment, an FMA-UE score of 20 to 50 and the presence of motor evoked potentials (MEPs) in a hand muscle following ipsilesional single-pulse TMS. Additional eligibility criteria are provided in the trial protocol (Supplement 1).

**Table 1.** Demographic and Clinical Characteristics of the 7 participants. Baseline demographic, lesion, and clinical characteristics for all participants (P1–P7). Stroke location was determined by clinical MRI and/or radiology report. Standard of care therapy (OT/PT) reflects weekly hours of occupational and physical therapy received during the 4-week baseline period, except where noted (P7: monthly session). *IS, ischemic stroke; ICH, intracerebral hemorrhage; FMA UE, Fugl-Meyer Assessment Upper Extremity; OT, occupational therapy; PT, physical therapy*.

| ID | Age range | Sex | Stroke type | Dominant Hand | Side Affected | Stroke Location | Time since stroke | Baseline FMA UE | Standard of Care |  |
| --- | --- | --- | --- | --- | --- | --- | --- | --- | --- | --- |
|  |  |  |  |  |  |  |  |  | OT | PT |
| P1 | 51-60 | F | IS | R | R | Subcortical | 3 years | 51 | - | 1 hr |
| P2 | 61-70 | M | IS | L | L | Subcortical | 1.5 years | 27 | - | - |
| P3 | 61-70 | M | IS | R | R | Mixed | 3 years | 41 | 2 hrs | - |
| P4 | 41-50 | M | IS | R | L | Mixed | 2 years | 43 | 0.75 hr | 1.5 hrs |
| P5 | 61-70 | F | IS | R | L | Subcortical | 8 years | 20 | 1 hr | - |
| P6 | 41-50 | F | ICH | R | R | Subcortical | 4 years | 27 | - | - |
| P7 | 51-60 | M | ICH | R | R | Mixed | 3 years | 30 | 1 hr/mo | - |

### Study Design and Intervention

Participants completed a 12-week protocol consisting of a 4-week standard-of-care baseline period, a 4-week intervention phase that included active kTMP combined with motor training (12 kTMP sessions; 3 sessions/week), and a 4-week post-intervention follow-up phase (see Supplement 1). All 7 participants completed the baseline and intervention phases, and 6 completed the final follow-up assessment. One participant withdrew prior to the Week 12 assessment. During the baseline period, 5 of 7 participants received concurrent physical and/or occupational therapy (see Table 1). These external rehabilitation therapies were discontinued at intervention onset. Safety and tolerability were monitored during each kTMP session.

### Clinical and Kinematic Assessments

The primary outcome was change in the FMA-UE at Weeks 0 (enrollment), 4 (pre-intervention), 8 (post-intervention), and 12 (follow-up). Clinical assessments were scored by an unblinded rater. As a secondary outcome, objective motor kinematics were collected during each intervention session using a Viper 16 motion tracking system (Polhemus Inc, Colchester, VT) while participants performed a standardized reach-to-grasp task. Movement quality was quantified using two kinematic metrics: spectral arc length (SPARC) ^10,11^, an index of movement smoothness in which higher (less negative) values indicate smoother movement trajectories (Figure 2A), and mean peak velocity count^12^, a measure of movement fragmentation in which fewer peaks reflect more ballistic, continuous movement trajectories (Figure 2B). Complete details regarding sensor acquisition and data processing are provided in the eMethods in the Supplement 2.

### Safety and Tolerability

At each session, participants were asked to report any device-related adverse reactions during and immediately following stimulation. Tolerability was also assessed using three questions adapted from prior TMS studies^13^ to evaluate annoyance, muscle twitches, and pain associated with stimulation, each rated on an 11-point scale (0 = no sensation, 10 = intolerable).

### kTMP Instrumentation and Targeting

The kTMP system (Magnetic Tides, Berkeley, CA) comprised a commercial high-current amplifier (AE Techron, Elkhart, IN) custom-modified to drive a MagVenture Cool-B70 coil (MagVenture A/S, Farum, Denmark). Using stereotaxic neuronavigation (Brainsight, Rogue Research Inc, Montreal, Quebec, Canada), the kTMP target was the ipsilesional motor cortex site that yielded the maximum MEP amplitude in the contralesional first dorsal interosseous muscle, using a previously described procedure^7^. The kTMP intervention used a 3.5-kHz carrier frequency with 3-Hz amplitude modulation, with stimulation parameters selected to produce cortical electric fields ∼7.5 V/m based on finite element method simulations in the standard SimNIBS “Ernie” head model^14^.

### Statistical Analysis

Due to the small sample size (N = 7), nonparametric methods were used. Changes in clinical outcomes across the four prespecified time points (Weeks 0, 4, 8, and 12) and changes in kinematic outcomes across the course of the kTMP intervention (pre-kTMP, 1 Week of kTMP, 4 Weeks of kTMP) were evaluated using a Friedman omnibus test, with effects reported using the Q statistic. Post hoc pairwise comparisons were performed using Wilcoxon signed-rank tests to evaluate specific within-participant transitions between time points. P values were adjusted for multiple comparisons using the Holm-Bonferroni correction, with statistical significance defined as α =.05. Statistical analyses were performed using Python version 3.10.18.

## Results

### Clinical Outcomes

Upper extremity motor impairment was tracked using the FMA-UE scale across the 12-week protocol. The omnibus Friedman test revealed a significant main effect of study period (*Q* = 16.12; *P* =.001). Mean FMA-UE scores remained stable during the 4-week standard-of-care baseline period, increasing from 34.1±11.0 (M ± SD) at enrollment to 34.3 ± 11.1 at the pre-intervention baseline (mean change, +0.14 points; 95% CI, –1.21 to 1.50; adjusted P =.98). Following the 4-week kTMP intervention (Week 8), mean FMA-UE scores increased to 41.3 ± 11.0, a mean increase of 7.00 points from the pre-intervention baseline (95% CI, 5.31 to 8.69; P-value = 0.016; adjusted for multiple comparisons P =.047; Figure 1C,1D). Six of the seven participants achieved the established minimal clinically important difference for upper extremity function in chronic stroke (MCID; ≥ 6-point improvement)^15^. At the 4-week post-intervention follow-up (Week 12), these gains were maintained in the 6 participants who completed this phase, with a mean FMA-UE score of 42.5 ± 8.96 and no significant decline from Week 8 (mean change, –1.33 points; 95% CI, –2.42 to –0.25; adjusted P =.13).

**Figure 1.**
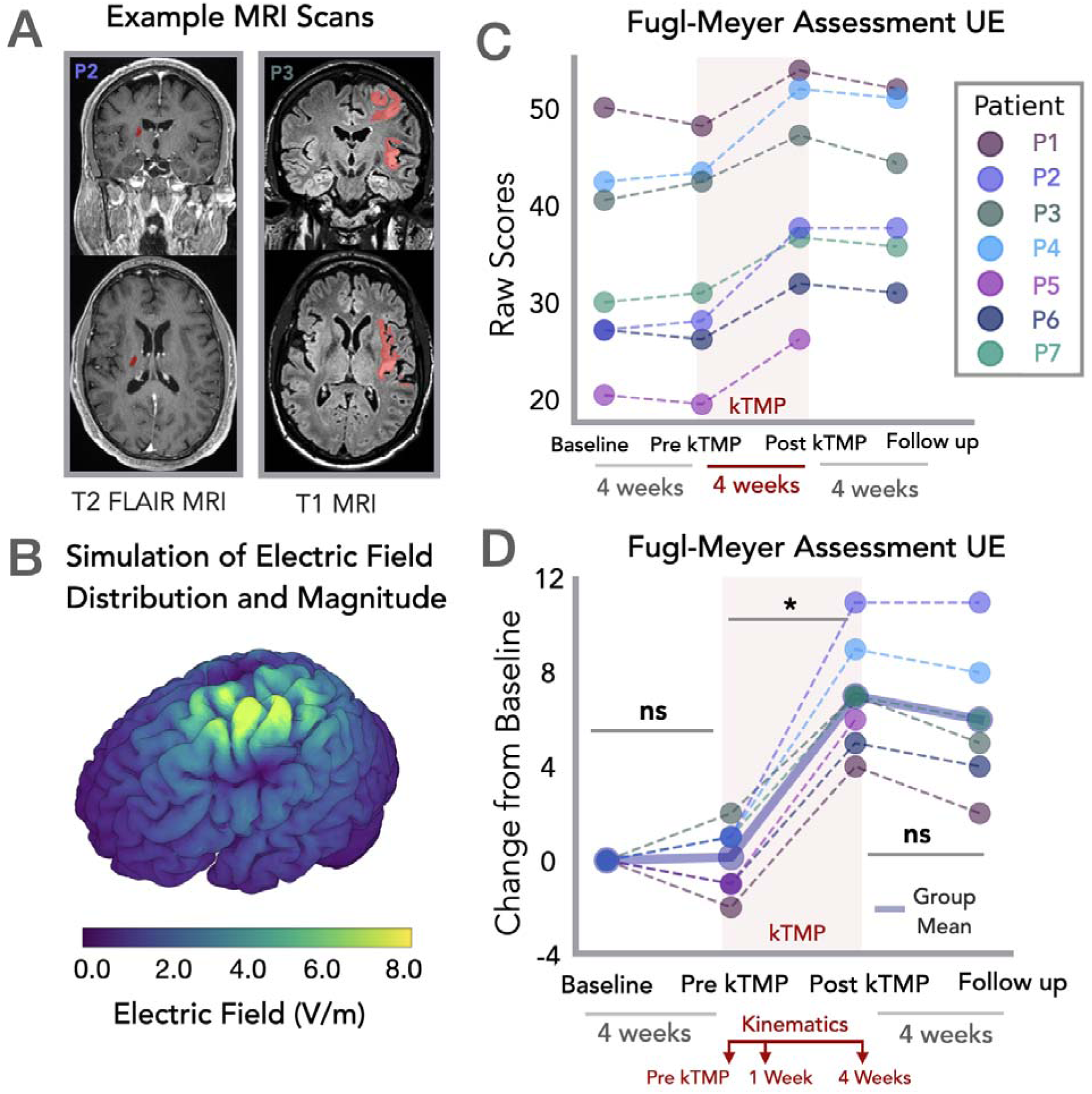
Lesion characterization, electric field modeling, and clinical motor outcomes following kTMP intervention. A. Representative MRI scans illustrating lesion location for two participants. Left: T2 FLAIR MRI (coronal and axial) for Participant 2 (P2); right: T1 MRI (coronal and axial) for Participant 3 (P3). Lesions are highlighted in red. B. Simulation of electric field distribution and magnitude for M1 target location using the standard head model in SimNIBS^14^. C. Raw Fugl-Meyer Assessment Upper Extremity (FMA-UE) scores for each participant (P1–P7) across the four assessment timepoints: Baseline, Pre-kTMP, Post-kTMP, and Follow-up. The shaded red region denotes the 4-week active intervention period. D. Change in FMA-UE scores from baseline for each participant, with the group mean overlaid (thick solid line). Scores remained stable during the 4-week baseline period (ns) and increased significantly following the 4-week kTMP intervention (*, adjusted P <.05) with gains largely sustained through the 4-week follow-up period (ns, Post-kTMP vs. Follow-up).

**Figure 2.**
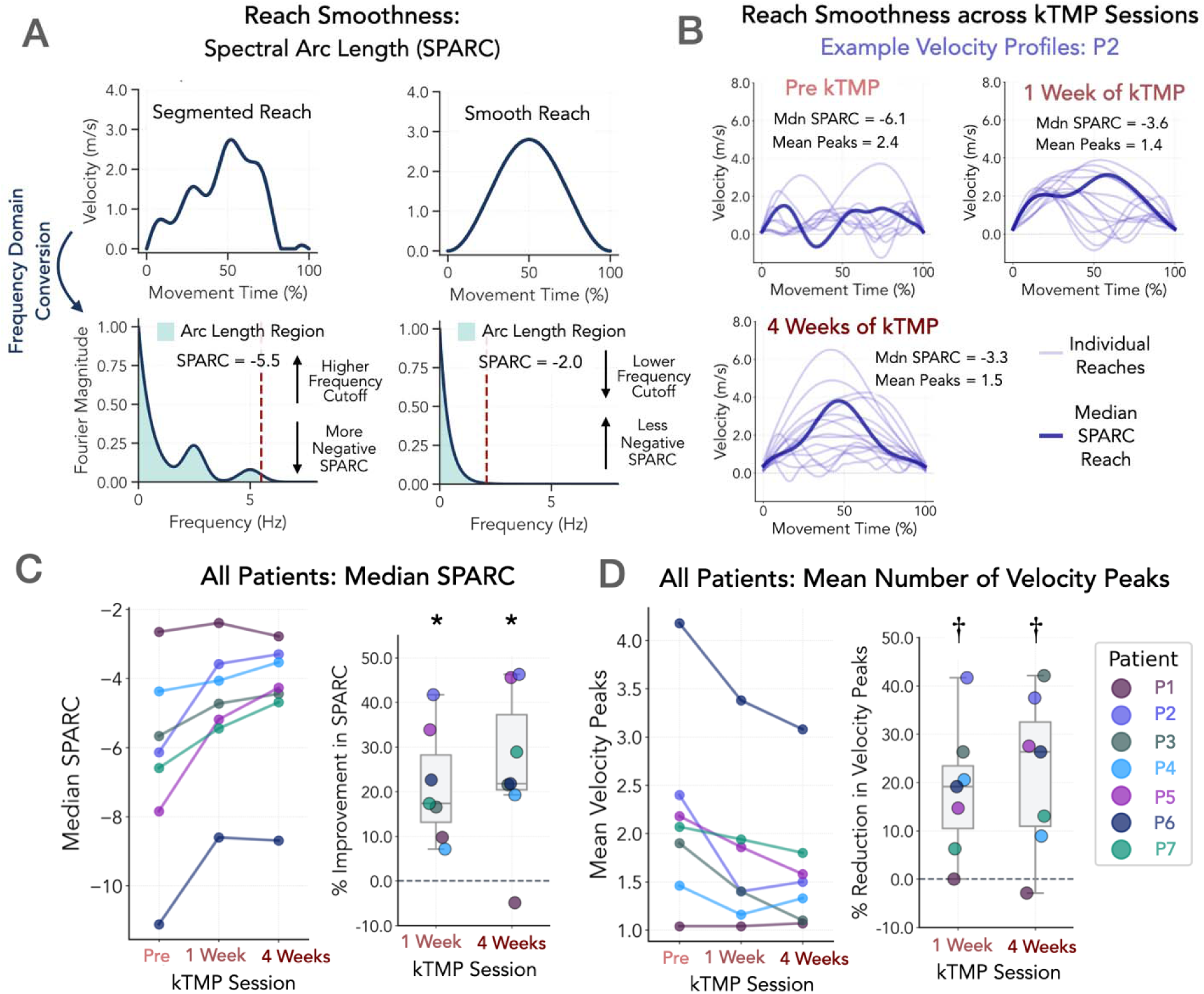
Kinematic assessment of reach smoothness and fragmentation across kTMP treatment. A. Schematic illustrating spectral arc length (SPARC) calculation from reach velocity profiles. Segmented (less smooth) reaches show a higher-frequency Fourier magnitude cutoff and correspondingly lower (more negative) SPARC values, while smooth reaches show a lower-frequency cutoff and less negative SPARC values. The cutoff frequency was defined as the point at which the normalized magnitude spectrum first fell below an amplitude threshold of 0.05^10^. B. Representative velocity profiles for an example participant (P2) across three timepoints: pre-kTMP (i.e., Day 0), 1 Week of kTMP, and 4 Weeks of kTMP. Thin lines represent individual reaches; thick lines represent the median reach for each timepoint. Median SPARC and mean number of velocity peaks are shown for each timepoint, illustrating a general increase in reach smoothness (less negative SPARC) and reduction in reach fragmentation (fewer velocity peaks) over the course of treatment (Velocity profiles for the remaining participants are shown in Supplementary Figure 2.). C. Left: Median SPARC values for all participants (P1–P7) at pre-kTMP, 1 Week, and 4 Weeks. Right: Percent improvement in SPARC from baseline at 1 Week and 4 Weeks (*, adjusted P <.05). D. Left: Mean number of velocity peaks for all participants across the same timepoints. Right: Percent reduction in velocity peak count from pre-kTMP at 1 Week and 4 weeks (†, nonsignificant trend after correction for multiple comparisons).

### Kinematic Outcomes

Upper extremity kinematics during a standardized reach-to-grasp task demonstrated significant improvements in movement smoothness over the 4-week intervention. The omnibus Friedman test revealed a significant effect of time (Day 0, 1 week of kTMP, 4 weeks of kTMP) for spectral arc length (SPARC; Q = 8.0; P =.018). Mean SPARC improved from –6.34 (2.68) at Day 0 to –4.86 (1.95) after 1 week of kTMP, a significant increase of 1.48 points (95% CI, 0.49 to 2.47; adjusted P =.03; Figure 2C). Improvements were maintained when measured at the end of Week 4 of kTMP, reaching –4.53 (1.96), for a total increase of 1.81 points from baseline (95% CI, 0.64 to 2. 98; adjusted P =.03).

For movement fragmentation, the omnibus Friedman test also demonstrated a significant effect of time for mean velocity peak count (Q = 6.74; P =.03). Mean peak count decreased from 2.18 (1.00) at Day 0 to 1.74 (0.80) at Week 1 (mean change, – 0.44 peaks; 95% CI, –0.77 to –0.10; adjusted P =.06) and 1.64 (0.69) at Week 4 (mean change, –0.54 peaks; 95% CI, –0.93 to –0.15; adjusted P =.06; Figure 2D). However, under Holm-Bonferroni correction, neither pairwise comparison reached statistical significance.

### Safety and tolerability

No device-related adverse events were reported. Tolerability ratings remained low throughout the 4-week intervention: annoyance (M = 0.14, range: 0.00–0.41), muscle activation (M = 0.03, range: 0.00–0.17), and pain (M = 0.05, range: 0.00–0.08), out of a possible 10.

## Discussion

This open-label pilot study provides preliminary evidence that noninvasive kTMP combined with motor training is feasible and may promote upper limb motor recovery in chronic stroke. Six out of seven participants exceeded the clinical MCID of 6 points, providing preliminary evidence of an intervention-related effect. Notably, the single participant who did not meet this 6-point threshold (P1) presented with the least severe impairment at baseline. Gains were sustained at a 4-week follow-up. Importantly, improvements in FMA-UE were accompanied by increased movement smoothness, providing an objective and continuous kinematic measure of motor recovery that captures dynamic aspects of movement quality not readily detected by the FMA, a more static clinical assessment.

### Limitations

This open-label, single-arm pilot study (n=7) lacked a sham-controlled group and FMA-UE scoring was performed by an unblinded assessor. A double-blind, randomized controlled trial with a larger sample is needed to confirm these findings and assess the possible contribution from the motor training and/or placebo effects. One participant withdrew before the Week 12 follow-up, resulting in missing data.

### Conclusion

These findings suggest that kTMP is a safe and well-tolerated intervention with the potential to facilitate upper extremity motor recovery in individuals with chronic stroke. These preliminary results provide rationale for larger, randomized controlled trials to further establish the efficacy of this novel, non-invasive neuromodulation approach.

## Data Availability

The data cannot be made publicly available upon publication because they contain sensitive personal information. The data that support the findings of this study are available upon reasonable request from the authors.

## Acknowledgements

This study was funded by a grant from the NIH and the Weill Neurohub.

## References

1. Hatem, S. M. et al. Rehabilitation of Motor Function after Stroke: A Multiple Systematic Review Focused on Techniques to Stimulate Upper Extremity Recovery. Frontiers in Human Neuroscience 10, (2016).

2. Kwakkel, G. & Dobkin, B. H. Vagus Nerve Stimulation for Upper Limb Function. Stroke 52, 3407–3409 (2021).

3. Liu, C. Y. et al. Vagus nerve stimulation paired with rehabilitation for stroke: Implantation experience from the VNS-REHAB trial. Journal of Clinical Neuroscience 105, 122–128 (2022).

4. Huang, Y. et al. Correction: Measurements and models of electric fields in thein vivohuman brain during transcranial electric stimulation. Elife 7, (2018).

5. Massimini, M., Peterchev, A. V., Vlachos, A. & Ziemann, U. Mechanisms of transcranial magnetic brain stimulation. Physiological Reviews 106, 2267–2316 (2026).

6. Nesmith, N. et al. Tactile Perception and Tolerability Thresholds of TMS Characterized by Intensity across Locations and Frequencies. 2026.01.28.26345065 Preprint at 10.64898/2026.01.28.26345065 (2026).

7. Labruna, L. et al. Kilohertz Transcranial Magnetic Perturbation (kTMP): A New Non-invasive Method to Modulate Cortical Excitability. eLife 13, (2024).

8. Khanna, P. et al. Low-frequency stimulation enhances ensemble co-firing and dexterity after stroke. Cell 184, 912–930.e20 (2021).

9. Merrick, C. M. et al. A systematic assessment of safety, tolerability and blinding effectiveness of kilohertz transcranial magnetic perturbation (kTMP). J. Neural Eng. 23, 044001 (2026).

10. Balasubramanian, S., Melendez-Calderon, A., Roby-Brami, A. & Burdet, E. On the analysis of movement smoothness. J Neuroeng Rehabil 12, 112 (2015).

11. Mohamed Refai, M. I., et al. Smoothness metrics for reaching performance after stroke. Part 1: which one to choose? J NeuroEngineering Rehabil 18, 154 (2021).

12. Rohrer, B. et al. Movement Smoothness Changes during Stroke Recovery. J. Neurosci. 22, 8297–8304 (2002).

13. Meteyard, L. & Holmes, N. P. TMS SMART - Scalp mapping of annoyance ratings and twitches caused by Transcranial Magnetic Stimulation. J. Neurosci. Methods 299, 34–44 (2018).

14. Saturnino, G., Antunes, A., Stelzer, J., & Thielscher, A. (2015). SimNIBS: A versatile toolbox for simulating fields generated by transcranial brain stimulation. Poster presented at 21st Annual Meeting of the Organization for Human Brain Mapping (OHBM 2015), Honolulu, HI, USA.

15. Page, S. J., Fulk, G. D. & Boyne, P. Clinically important differences for the upper-extremity Fugl-Meyer Scale in people with minimal to moderate impairment due to chronic stroke. Phys Ther 92, 791–798 (2012).

