## Supplementary Document 1 for "Kilohertz Transcranial Magnetic Perturbation (kTMP) for Upper Extremity Motor Recovery After Stroke: A Nonrandomized Pilot Trial"

### Supplement 1: Pilot kTMP Intervention Protocol

#### Overall Study Design

Participants completed a 12-week protocol consisting of a 4-week standard-of-care baseline period, a 4-week intervention phase, and a 4-week post-intervention follow-up phase. This is an open-label study; all participants received active kTMP (no sham condition) during the intervention phase.

Of the 7 enrolled participants, all completed the baseline and intervention phases and 6 completed the Week 12 follow-up assessment. One participant withdrew prior to the final assessment.

Concurrent rehabilitation therapy: During the baseline period, 5 of 7 participants were receiving concurrent physical and/or occupational therapy. These external rehabilitation therapies were discontinued at intervention onset to isolate the effects of kTMP combined with motor training.

#### Intervention Phase: Session Structure

Each of the 12 treatment sessions lasted approximately 90–120 minutes and included three components: setup/preparation, kTMP delivery paired with motor tasks, and post-stimulation consolidation.

1. Setup and Preparation (10–20 minutes)

- Participant positioning for comfort and consistency across sessions.
- kTMP coil placement using stereotaxic neuronavigation (Brainsight, Rogue Research Inc., Montreal, Quebec, Canada) to target the ipsilesional primary motor cortex site yielding the maximum MEP amplitude in the contralesional first dorsal interosseous (FDI), using a previously described procedure (Ref. 6). This hotspot was defined during screening/baseline.
- Verification of coil orientation and stability prior to stimulation.

Instrumentation: The kTMP system (Magnetic Tides, Berkeley, CA) comprised of a commercial high-current amplifier (AE Techron, Elkhart, IN) custom-modified to drive a MagVenture Cool-B70 coil (MagVenture A/S, Farum, Denmark).

2. kTMP Delivery Paired with Motor Tasks (~60 minutes)

Stimulation parameters: The intervention used a 3.5 kHz carrier frequency with 3 Hz amplitude modulation, with stimulation parameters selected to produce cortical electric fields close to 7.5 V/m based on the SimNIBS "Ernie" template brain mode (Saturnino et al. 2015).

Protocol note: As this was a pilot study, the task paired with stimulation was refined during the course of the trial. The original design called for functional motor tasks to be performed concurrently with kTMP delivery. However, we observed that participants with more severe impairment produced significant head movement during the functional tasks which compromised coil-to-target alignment and, consequently, stimulation delivery. The protocol was therefore adapted as follows:

- P1–P3: Functional tasks performed concurrently with kTMP stimulation.
- P4–P7: Simple, well-controlled motor tasks performed concurrently with kTMP stimulation (to maintain head/coil stability), followed immediately by functional tasks performed after stimulation ended.

Simple, well-controlled motor tasks (performed during kTMP in P4–P7): Isolated movements designed to activate the ipsilesional motor network without inducing compensatory head movement, including:

- Finger abduction/adduction
- Wrist flexion/extension
- Grip-and-release

Functional tasks (performed during kTMP in P1–P3; performed immediately post-stimulation in P4–P7):

- Reach and Grasp – reaching for and grasping objects of varying sizes (cricket ball or marble)
- Gross Movement – large-amplitude arm movements (reaching, lifting, pushing/pulling)
- Object Flipping – manipulating and reorienting objects in hand
- Simulated Eating Tasks – fork/spoon use, cup lifting, simulated meal preparation
- Opening Containers – unscrewing lids, opening boxes, operating latches

3. Post-stimulation tolerability assessment

Immediately following stimulation, participants were asked three questions to assess stimulation-related sensations and tolerability, adapted from the framing used in prior TMS studies (Ref. 11):

1. "Did you experience any annoyance from the stimulation?"
2. "Did you experience any muscle twitches from the stimulation?"
3. "Did you experience any pain from the stimulation?"

Each item was rated on an 11-point scale (0 = no sensation; 2 = just noticeable; 4 = noticeable but not aversive; 6 = somewhat uncomfortable; 8 = very uncomfortable; 10 = intolerable).

For the muscle twitch item, participants were instructed that this may include incidental muscle activation in the scalp, face, or neck, as well as twitches in the contralateral body when stimulating over motor cortex.

4. Post-Stimulation Consolidation (10 minutes)

- Continued functional task practice to consolidate gains from the stimulation-enhanced learning period

**Clinical and Kinematic Assessments**

Primary outcome: Change in upper extremity motor impairment, assessed using the Fugl-Meyer Assessment–Upper Extremity (FMA-UE) at Weeks 0 (enrollment), 4 (pre-intervention), 8 (post-intervention), and 12 (follow-up). The FMA-UE is graded from 0 (hemiplegia) to 66; higher scores indicate better motor control. Clinical assessments were video-recorded and scored by an unblinded rater.

Secondary outcome: Objective motor kinematics, collected during each intervention session using a Viper 16 electromagnetic motion tracking system (Polhemus Inc., Colchester, VT) while participants performed a standardized reach-to-grasp task. Movement quality was quantified using:

- Spectral Arc Length (SPARC): a frequency-domain index of movement smoothness; higher (less negative) values indicate smoother movement trajectories
- Mean velocity peak count: a measure of movement fragmentation; fewer peaks reflect more ballistic, continuous movement trajectories

**Standardized reach-to-grasp task**

Participants were directed to a starting position on the table, marked with a red X. From this position, participants reached 10 cm toward a target object mounted on a vertical rod fitted with an electromagnetic sensor, grasped the object, lifted it 5 cm, with upward movement constrained by a stop ring around the rod, and finally lowered the object to return to the starting position.

*Object selection:* All participants initially performed the task with the cricket ball. Participants who achieved at least 8 of 10 successful trials with the cricket ball were subsequently tested with the marble. Those who achieved at least 3 of 10 successful trials with the marble continued with the marble for the remainder of the study; those who did not meet this threshold continued with the cricket ball. This staged selection procedure resulted in the following object assignments:

- Cricket ball: P2, P4, P5, P6, P7
- Marble: P1, P3

Complete details regarding sensor acquisition and kinematic data processing are provided in the separate kinematic analysis supplement.

**Safety and Tolerability Monitoring**

In addition to completing the tolerability questionnaire described above, participants were asked during each session to report any device-related adverse reactions occurring during or immediately following stimulation.

**Inclusion Criteria**

1. Age 18–80 years with stable upper limb motor dysfunction caused by a single ischemic and/or hemorrhagic stroke occurring more than 6 months prior to recruitment
2. Fugl-Meyer Assessment: Upper Extremity (FMA-UE) score of 20–50
3. Presence of motor evoked potentials (MEPs) in a hand muscle upon stimulation of ipsilesional cortex with suprathreshold single-pulse TMS (MEP+), used as a proxy of residual corticospinal function

**Exclusion Criteria**

1. Cognitive/Language

- Cognitive impairment (MoCA < 20)
- Language impairment that interferes with the ability to adhere to the protocol or provide informed consent

2. Medical

- Pregnancy or plans to become pregnant
- Uncontrolled medical conditions, including but not limited to severe cardiovascular or cardiopulmonary disease
- Severe alcohol or drug abuse within the past year
- Major depression

3. Contraindications to Non-Invasive Brain Stimulation

- History of seizures

- History of epilepsy or family history of epilepsy
- History of migraines or other severe headaches
- Permanent eyeliner (tattooed)
- Presence of metal implants in the head (excluding the mouth)
- Implanted electronic devices
- High blood pressure not controlled by medications
- Hearing loss or hearing impairment
- Chronic or transient sleep disturbance (e.g., jet lag)
- History of fainting
- Chronic stress-related conditions
- Pregnancy or plans to become pregnant during the trial
