## Supplementary Document 2 for "Kilohertz Transcranial Magnetic Perturbation (kTMP) for Upper Extremity Motor Recovery After Stroke: A Nonrandomized Pilot Trial"

### Kinematic Outcome Measures

Hand kinematics during the reach phase of the reach-to-grasp task were recorded using an electromagnetic motion capture system (Polhemus ViperX 16, sample rate = 240 Hz). A sensor was placed on the palm, providing 3-D measurements of hand position. The raw position data were smoothed using a fourth-order zero-phase Butterworth low-pass filter with a 6 Hz cutoff frequency, implemented using forward-backward filtering to prevent phase distortion. Velocity profiles were computed by numerical differentiation of the filtered position time series.

#### *Reach Segmentation*

Individual reach trials were identified from the continuous kinematic record using a velocity-based segmentation algorithm applied to the dominant movement axis, defined as the dimension of greatest positional displacement across the pre-kTMP recording session. Candidate reach trials were validated using displacement criteria requiring a minimum displacement in the primary movement axis, which ranged from 20–45% of the full session positional range across patients. Trials were also required to have a duration between 0.2 and 10 seconds. Overlapping duplicate segments were removed by retaining the longer of any two segments sharing more than 80% overlap.

To optimize the number of recovered trials, we opted to individualize segmentation parameters given the substantial between-subject variability inherent in clinical stroke populations as well as differences in task execution across participants.  To identify movement onset, we first identified the time of peak velocity. Moving backwards, movement onset was defined as the last sample in which velocity remained below a threshold for a sustained period. The threshold was set to 5-10% of peak velocity for that movement and the required sustained duration ranged from 100-200 ms. Movement offset was defined in a similar manner, the first point following peak velocity at which velocity remained below the same threshold for a sustained period. The parameters remained fixed across all sessions for a given patient, ensuring longitudinal comparability.

*Number of Velocity Peaks*

Movement fragmentation was quantified by counting the number of discrete velocity peaks within each segmented reach velocity profile. Peaks were identified using a prominence-based detection algorithm applied to the velocity time series. To avoid detecting spurious peaks arising from signal noise while remaining sensitive to genuine submovements, two thresholds were applied. First, a minimum prominence threshold was set at 15% of that trial's peak velocity, requiring each detected peak to rise sufficiently above its surrounding local minima to be counted as a distinct submovement (Kahn et al., 2001). Second, a minimum inter-peak distance of 10% of the total trial duration was enforced to prevent adjacent samples of the same submovement from being counted as separate peaks. Both thresholds were scaled to each individual trial rather than set as fixed absolute values, ensuring consistent sensitivity despite the considerable variability in movement speed and reach duration. The mean number of peaks across trials was computed within each session to produce a session-level estimate, with a value of 1 indicating a single smooth bell-shaped velocity profile consistent with uninterrupted voluntary reaching.

*Movement Smoothness (SPARC)*

Movement smoothness was quantified using the Spectral Arc Length (SPARC) metric (Balasubramanian et al., 2015). SPARC measures the arc length of the normalized magnitude spectrum of the velocity profile, with more negative values indicating less smooth, more fragmented movement. Prior to computation, velocity profiles were resampled to a fixed duration of 240 samples (equivalent to 1 second at 240 Hz) to ensure comparability across trials and sessions with variable movement durations, consistent with the reference duration used in normative reach-to-grasp data (Vlaar et al., 2021). Reaches shorter than 300 ms were excluded from SPARC computation given that such trials produce unreliable spectral estimates. The magnitude spectrum was computed using a zero-padded fast Fourier transform (padding factor = 4).

The cutoff frequency for arc length computation was defined as the frequency at which the normalized magnitude spectrum first fell below an amplitude threshold of 0.05, consistent with the parameters recommended in the original SPARC validation (Balasubramanian et al., 2015). The maximum cutoff frequency was set at 15 Hz. This exceeds the recommended 10 Hz (20π rad/s) cutoff, accommodating one participant with severe motor impairment whose velocity spectrum contained meaningful energy only up to approximately 11 Hz. Increasing the maximum cutoff frequency for this participant ensured that the full spectral content of their highly fragmented movement was captured rather than artificially truncated.

Velocity profiles were normalized to a positive peak of 1 prior to spectral analysis, with negative velocity values set to zero to prevent spurious low-frequency spectral content from movement reversals. Trial-level SPARC values were averaged within each session to produce a session-level estimate.

Balasubramanian, S., Melendez-Calderon, A., Roby-Brami, A., & Burdet, E. (2015). On the analysis of movement smoothness. *Journal of NeuroEngineering and Rehabilitation*, *12*, 112. https://doi.org/10.1186/s12984-015-0090-9

Kahn, L. E., Zygman, M. L., Rymer, W. Z., & Reinkensmeyer, D. J. (2001). Effect of robot-assisted and unassisted exercise on functional reaching in chronic hemiparesis. *2001 Conference Proceedings of the 23rd Annual International Conference of the IEEE Engineering in Medicine and Biology Society*, *2*, 1344–1347 vol.2. https://doi.org/10.1109/IEMBS.2001.1020445
