## Supplementary Figure 1 for "Kilohertz Transcranial Magnetic Perturbation (kTMP) for Upper Extremity Motor Recovery After Stroke: A Nonrandomized Pilot Trial"

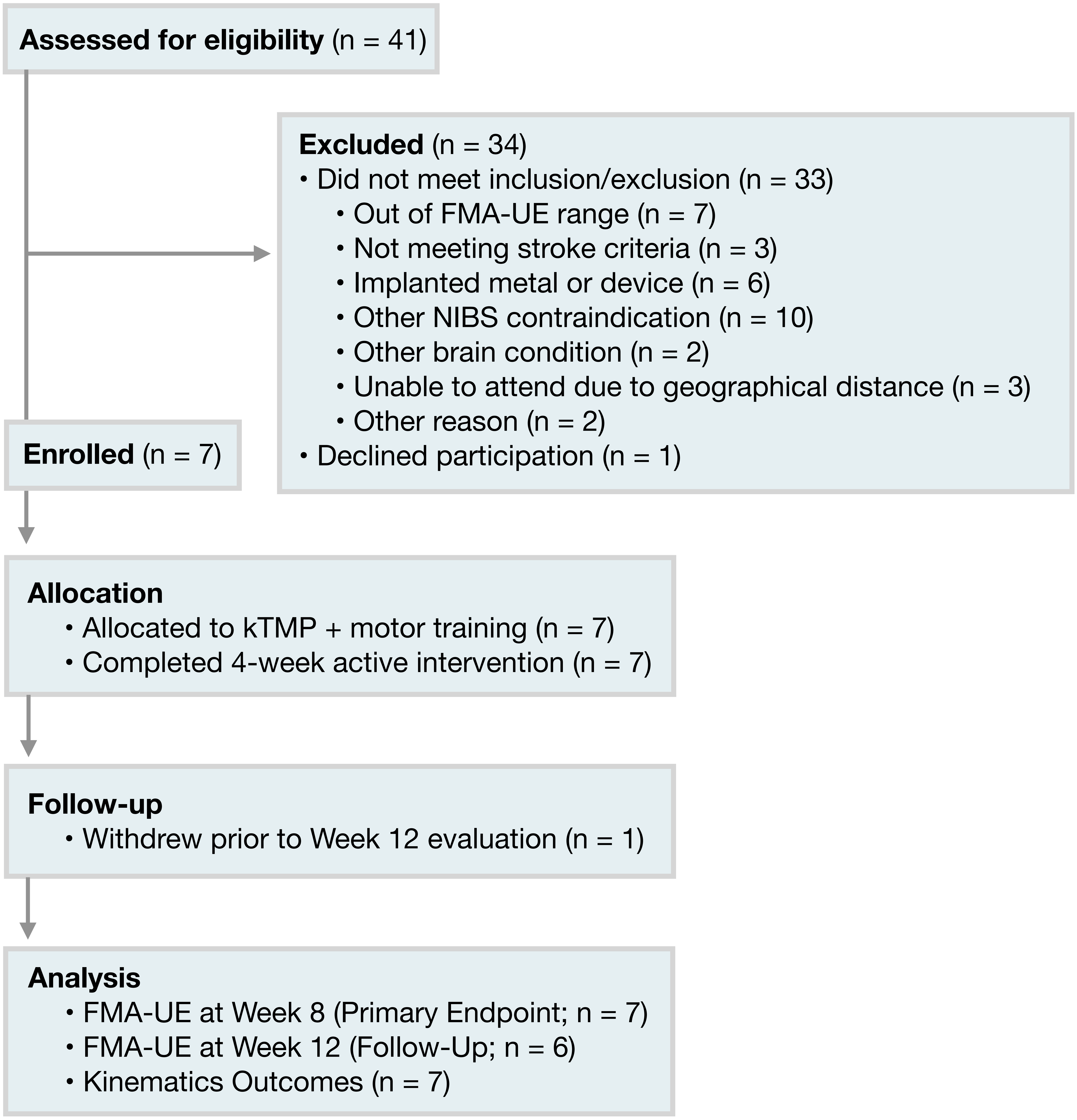

Assessed for eligibility (n = 41)
Excluded (n = 34)
• Did not meet inclusion/exclusion (n = 33)
• Out of FMA-UE range (n = 7)
• Not meeting stroke criteria (n = 3)
• Implanted metal or device (n = 6)
• Other NIBS contraindication (n = 10)
• Other brain condition (n = 2)
• Unable to attend due to geographical distance (n = 3)
• Other reason (n = 2)
• Declined participation (n = 1)
Enrolled (n = 7)
Allocation
• Allocated to kTMP + motor training (n = 7)
• Completed 4-week active intervention (n = 7)
Follow-up
• Withdrew prior to Week 12 evaluation (n = 1)
Analysis
• FMA-UE at Week 8 (Primary Endpoint; n = 7)
• FMA-UE at Week 12 (Follow-Up; n = 6)
• Kinematics Outcomes (n = 7)

**Supplement figure 1**. Study flow diagram. Forty-one individuals were screened, leading to the enrollment of 7 participants in the study. All the participants completed the 4-week active intervention and were included in the primary endpoint (FMA-UE at Week 8) and kinematic analyses. One participant declined to return for the follow-up, leaving 6 participants who were tested at Week 12.
