## Supplementary Figure 2 for "Kilohertz Transcranial Magnetic Perturbation (kTMP) for Upper Extremity Motor Recovery After Stroke: A Nonrandomized Pilot Trial"

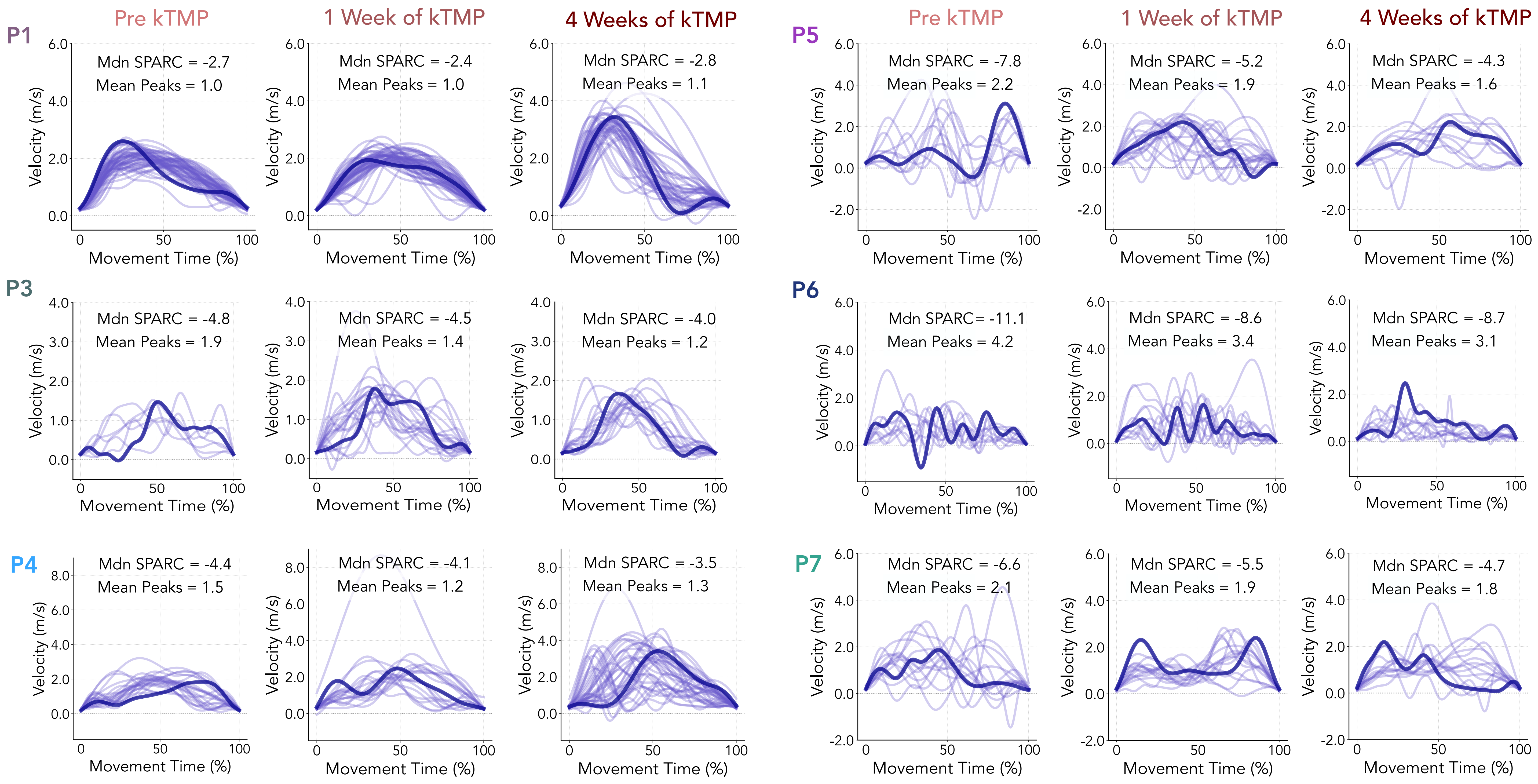

Pre kTMP
1 Week of kTMP
4 Weeks of kTMP
P1
Mdn SPARC = -2.7
Mean Peaks = 1.0
Mdn SPARC = -2.4
Mean Peaks = 1.0
Mdn SPARC = -2.8
Mean Peaks = 1.1
P5
Mdn SPARC = -7.8
Mean Peaks = 2.2
Mdn SPARC = -5.2
Mean Peaks = 1.9
Mdn SPARC = -4.3
Mean Peaks = 1.6
P3
Mdn SPARC = -4.8
Mean Peaks = 1.9
Mdn SPARC = -4.5
Mean Peaks = 1.4
Mdn SPARC = -4.0
Mean Peaks = 1.2
P6
Mdn SPARC= -11.1
Mean Peaks = 4.2
Mdn SPARC = -8.6
Mean Peaks = 3.4
Mdn SPARC = -8.7
Mean Peaks = 3.1
P4
Mdn SPARC = -4.4
Mean Peaks = 1.5
Mdn SPARC = -4.1
Mean Peaks = 1.2
Mdn SPARC = -3.5
Mean Peaks = 1.3
P7
Mdn SPARC = -6.6
Mean Peaks = 2.1
Mdn SPARC = -5.5
Mean Peaks = 1.9
Mdn SPARC = -4.7
Mean Peaks = 1.8
Velocity (m/s)
Movement Time (%)

**Supplement figure 2.** Velocity profiles for the other six participants following the same format as in Figure 2b: pre-kTMP (i.e., Day 0), 1 Week of kTMP, and 4 Weeks of kTMP. Thin lines represent individual reaches; thick lines represent the median spectral arc length (SPARC) reach for each timepoint. Median SPARC and mean number of velocity peaks are shown for each timepoint, illustrating a general increase in reach smoothness (less negative SPARC) and reduction in reach fragmentation (fewer velocity peaks) over the course of treatment.
